# An evolving case for centering health equity as a research priority in predominantly White academic medical centers

**DOI:** 10.1101/2024.10.02.24314778

**Authors:** Elizabeth A. Bonney, Gagan Deep Bajaj, Olivia O. Darko, Maria Mercedes Avila, Brittany M. Williams

## Abstract

**Purpose:** Health disparities impact those minoritized by race, class, gender nonconformance, and rurality. There is an urgent need to shift scholarly focus from merely documenting health disparities to effecting change through health equity research. The present mixed-methods study explores the motivation, opportunities, and barriers in performing health equity research in a majority white college of medicine.

**Methods:** We use landscape analysis: surveys, focus groups, and interviews with expressly interested faculty. Results: Our findings suggest there are barriers to communication, access, and recognition for health equity research that impact and influence the existence and possibilities of health equity scholarship in Vermont.

**Conclusion:** Colleges of medicine are increasingly recognized as being responsible for advancing Health Equity. Our findings underscore the necessity of this work within the state of Vermont and offer recommendations to remove barriers at our institution and others similarly situated.

## BACKGROUND AND INTRODUCTION

The healthcare system has played a vital role in slavery^1^, forced sterilization^2^, providing the intellectual basis for discrimination, inappropriate clinical trials^3^ unethical medical experimentation, and lack of intervention against group-targeted societal violence^4^. This history governs the basis of the societal issues affecting our current healthcare system, and further contributes to factors that have eroded trust between healthcare providers, marginalized patients and their communities. For example, Black and Hispanic patients are more likely to report distrust in their healthcare providers compared to their white peers^5^, which might contribute to poor health and mental healthcare outcomes in these patients and communities. Research suggests a complex interaction between different variables, including socioeconomic factors, city of residence, and race/ ethnicity and provider distrust^6^.

Solving the complexities of marginalized group health requires approaches beyond superficially recognizing differences in disease outcomes. Scholars must use a holistic lens to shift from a focus on health disparity towards a health equity^7^. This requires contextualization beyond access to care^8,9^ and broad examination of health system-related factors including the diversity of the healthcare workforce and disease destigmatization^10^.

Provider groups differ in their belief that health disparities exist and are based on income, English literacy, education, or race/ethnicity^11^. White and Asian physicians are less likely to acknowledge healthcare disparities compared to physicians of other groups: most believe that health insurance status is the principal driver of healthcare disparities^11^, suggesting that while they understand potential gaps in healthcare access, they have yet to connect this to systemic causes, which cannot be corrected by provider education alone. Evidence suggests that when healthcare providers are educated on bias and its role in perpetuating health disparities, generated changes in bias awareness are often not sustained^12^. This waning awareness occurs concurrently with structural and systemic forces that maintain health inequities^12^ and further does not fully explain why individual providers retain prejudices against marginalized communities that negatively impact their care. Another explanation can be found in the very nature of academia itself: who gets to participate, who is supported and meaningfully heard, whose trauma is adequately addressed, who does work that is valued^13^, and who gets to lead^14^. Black women across academe have (in)formally asked these questions^15^.

Yet, the onus of asking and answering questions relevant to health equity cannot rest squarely on those marginalized in the medical profession. Research not only into health disparities, but also health equity must be seen as vital to systematically enhancing the health of the entirety of medicine. Health equity research is the critical route by which we derive, test, and assess evidence-based interventions to alleviate health inequities and uplift and center marginalized communities. It encompasses all critical methods and approaches to intentional, disciplined and scientific inquiry that could be harnessed in achieving true health equity. Such research must also substantively involve healthcare consumers and communities—making no conclusions without consulting their members. If done appropriately such work is a mechanism to acknowledge and interrogate historical wrongs and heal relationships with the healthcare system^16,17^. It must ensure community access to information, opportunities for project oversight, authorship and training, and pathways to incorporate change^18^.

Despite the existence of organizationally diverse groups that are actively engaged in health equity research, many academic environments persist in a state of relative underdevelopment in this area^19^. We believe Vermont to be one of those environments. Here, historically unserved and underserved communities, including former refugee, immigrant, aboriginal, migrant farm-working, and Hispanic/Latinx communities have experienced negative interactions with healthcare centers, providers and researchers^5,6^. This discord has a historical context. For example, Vermont supported forced sterilization of indigenous peoples^20^ and lagged in its formal recognition of members of this group.

We sought to determine Vermont’s academic medical environment and its trajectory in elevating health equity as one of its strategic priorities. Through an examination of publication data, surveys, focus groups and interviews we sought to answer the specific questions: (1) what is the current landscape of health equity research in and on the State of Vermont; and (2) how do scholars who espouse a commitment towards diversity, equity, and inclusion in Vermont understand and operationalize health equity? Our findings suggest that Vermont is ripe for further investment in mentorship, collaboration, and innovation in health equity scholarship.

## METHODS

### Human subjects

The Research Protections Office of the University of Vermont deemed the project to be “not human-subjects research”.

### Literature search

We searched English language literature from medicine, science, and health-related databases (Medline Ovid, World of Science, and CINHAL) using the terms health disparities, health equity, health equity research, social justice, and Vermont. We read the citation titles/abstracts for relevance and catalogued author affiliations. Six searches were done including two per database: one for Vermont-affiliated publications; the second for all publications. Searches were conducted June 23rd, 2022. Each probe was time-limited to January 1, 2000 to December 31, 2021 (few Vermont-related articles met criteria before 2000). Our refined Web of Science search included citations related to medicine and public health from the “Science Citation Index Expanded” collection. We imported chosen citations into EndNote, removed duplicates and removed non-peer-reviewed article citations. After cleaning, citations were exported to Microsoft Excel for analysis. Two investigators (Authors 1 and 3) reviewed the list of VT-affiliated articles and removed those deemed irrelevant.

A list of scholars in collaborating institutions was used to further validate the articles.

### Interest in pursuing Heath Equity research at the Larner College of medicine (LCOM)

#### 1) Survey

To gauge faculty interest in health equity research, we connected with the developing administrative network driven by the Office of Diversity Equity and Inclusion within LCOM. This office supports “diversity champions” for each department. A survey was sent out to diversity champions and other key stakeholders who had participated in the 2019 cohort of the Diversity, Equity and Inclusion Certificate program at LCOM. The respondents were from all career stages within LCOM. Interested participants were invited back for focus groups.

#### 2) Faculty focus groups

To further understand interest in health equity research we conducted four online, focus group sessions^21^ from December 14, 2020 to February 11, 2021. Each 1.5-hour focus group session comprised up to 8 participants, and included at least one facilitator. Participants (15) were from eight academic units. Twelve identified as women or gender minorities, 3 identified as male, 14 as white, and one of color.

Discussion built on questions similar to those in the diversity champion survey. All participants received a summary of orienting definitions, purpose, questions to which to respond in advance. Definitions used were scholarly-based composites that synthesized the current landscape and best practices^22^ The sessions were recorded and transcribed later for analysis. We analyzed the group findings for the presence of themes as they spontaneously evolved^21^.

## RESULTS

### Trends in publications focused on health equity research in Vermont

**Figure 1** shows encouraging trends in publications in Health Equity from Vermont (**Figure 1B**). We estimate that over 80% of the 56,297 English language journal articles published from 2000 to 2021 found using our search criteria were critically relevant to aspects of health equity. We also found and manually verified that 252 of these were published in affiliation with VT or UVM. Despite the significant difference in the volume of VT-affiliated journal articles in comparison to all English articles **(Figure 1A)**, both analyses demonstrate a notable increase in cumulative publications following 2012. ***This suggests a possible push for health equity research over the past 10 years***. We also compiled a list of investigators in or affiliated with Vermont academic or health-related institutions such as the Vermont Department of Health and the VA Medical Center at White River Junction. We estimate that approximately 60 published investigators related to Vermont institutions are engaged or might be interested in health equity research.

**Figure 1:**
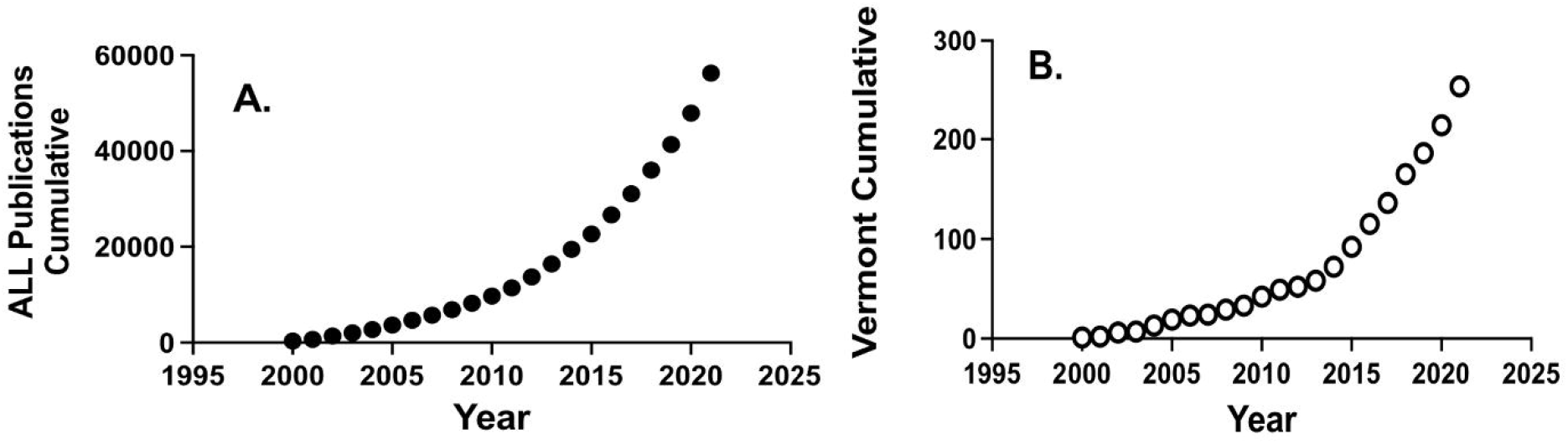
Comparison of cumulative publications relevant to Health disparities and Health equity in Vermont. **1A:** Analysis of all publications through 2021. **1B:** Vermont-associated publications.

### Survey of Diversity Champions

We invited all LCOM departmental diversity champions and participants in a recent diversity training program to participate in a survey. Of the 31 invited participants, 17 responded, and 15 (48.4%) completed this survey. 73% of the respondents were interested in performing health disparity/equity research, while 47% indicated departmental support for this work. 46% of respondents identified other colleagues potentially working or interested in this work. From the responses, we identified names of 12 additional faculty to invite for the focus groups. In all, we invited 19 faculty to participate in the focus groups.

### Faculty focus groups

Fifteen faculty (12 women and 3 men) from 8 academic departments participated, and were mostly white-identified.

We used a conversational style that facilitated dissemination of information, including establishing definitions^22^. In addition to definitions, the group discussed feasibility, barriers and opportunities related to doing this work for a career (**Table 1**).

**Table 1.**
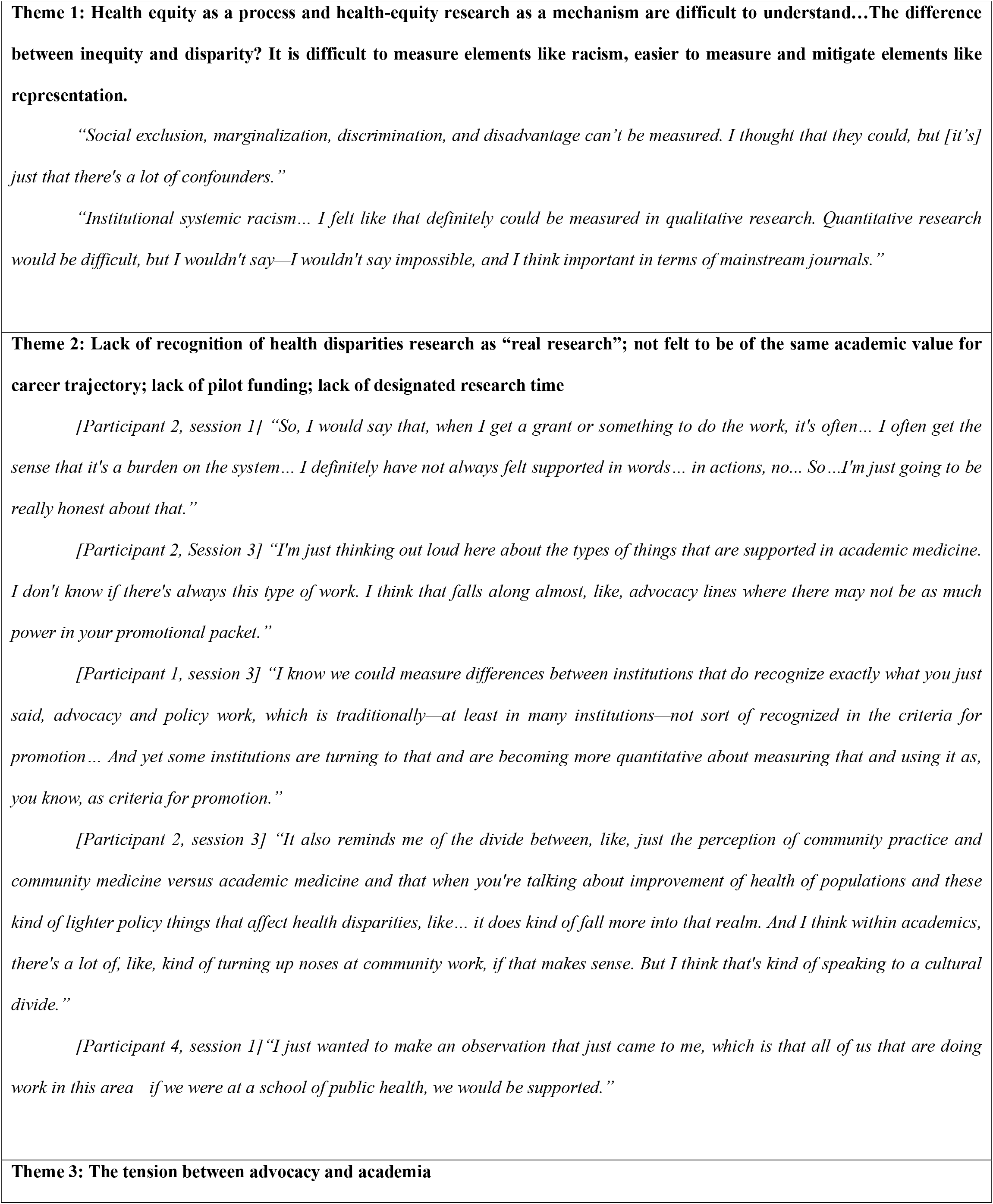
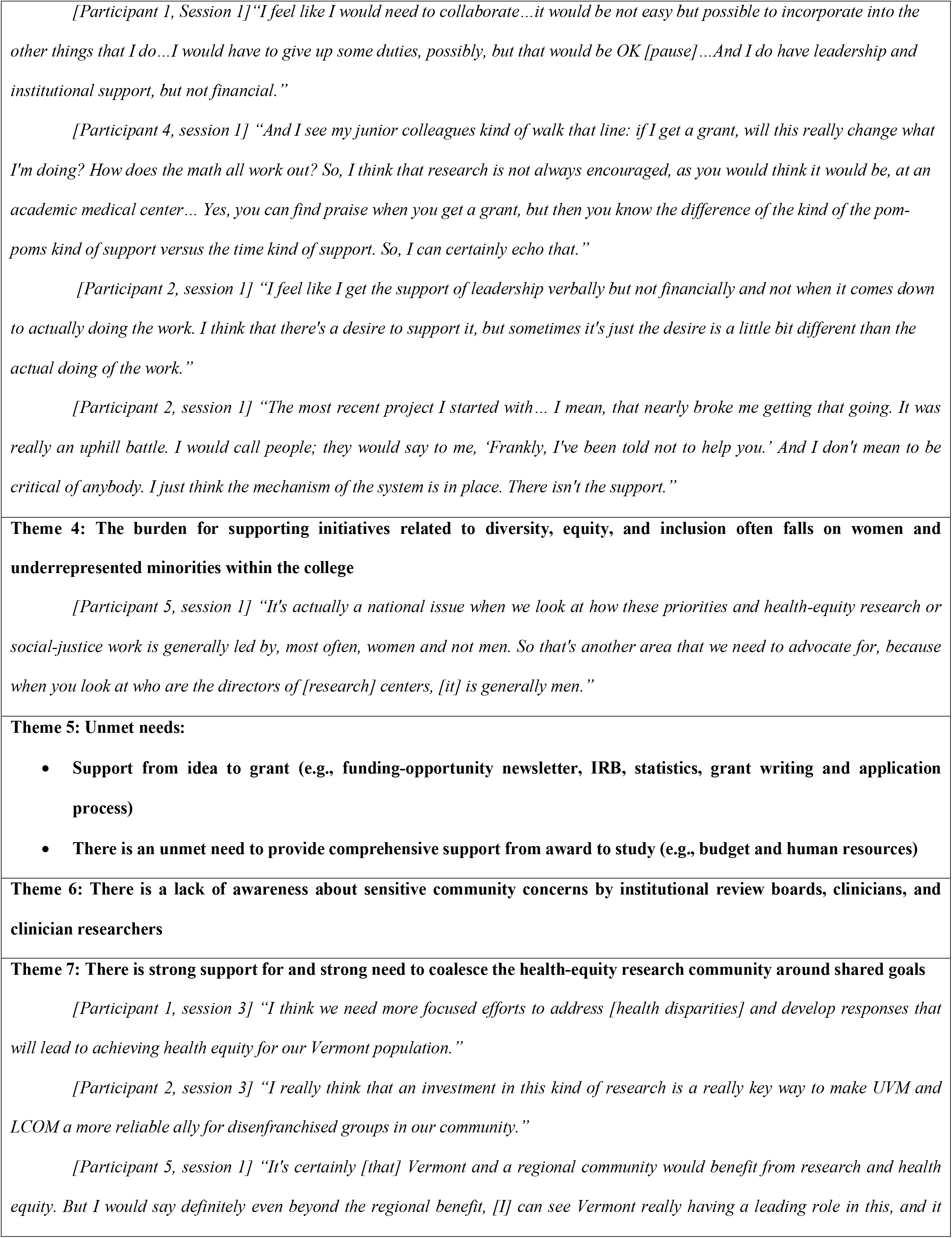
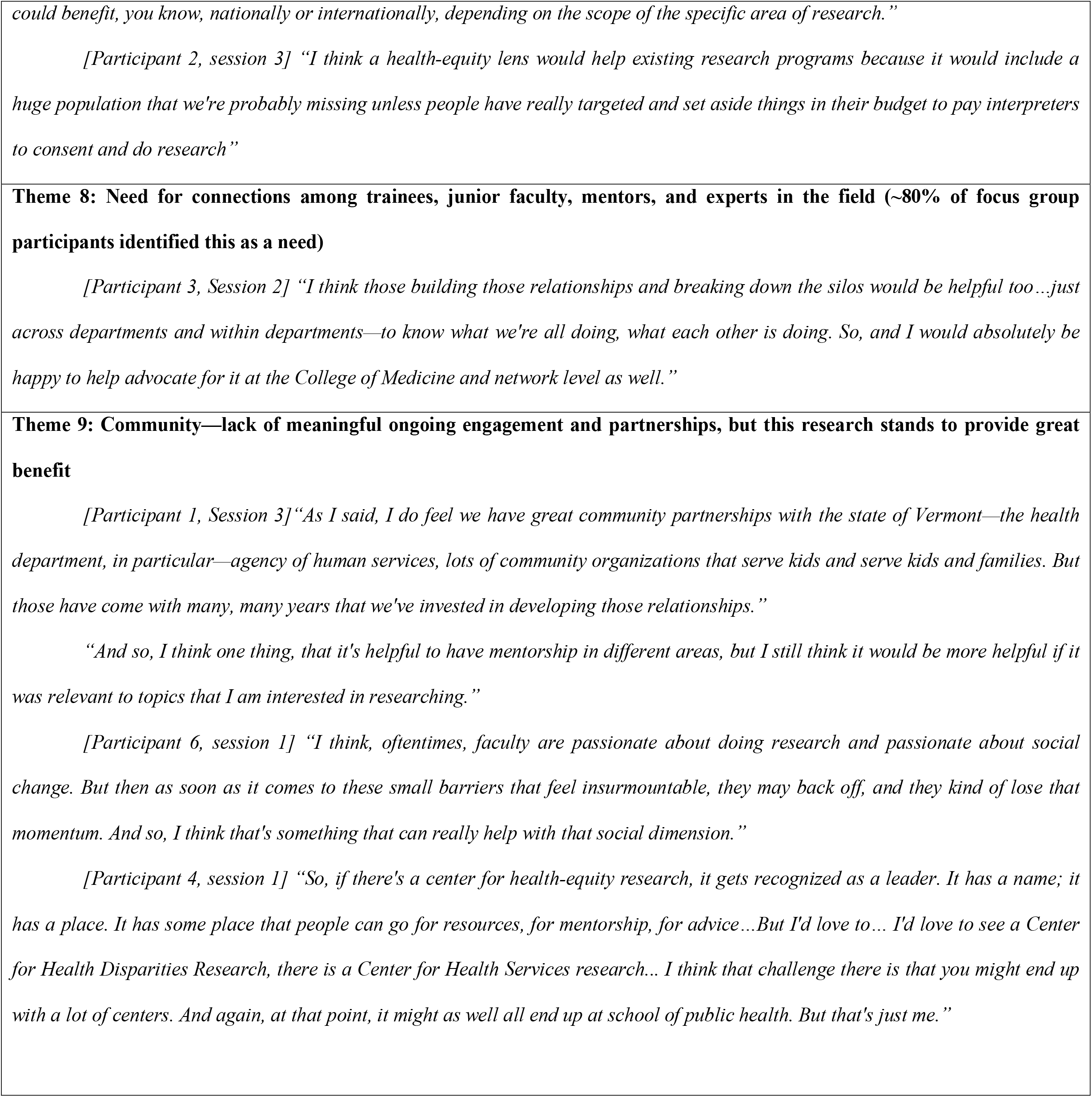
Themes that emerged during faculty focus groups’ discussions.

Many participants were concerned about the definitions of health disparity and health equity. Moreover, some participants were more willing to attribute lack of healthcare access to rurality and age rather than categorize barriers as mechanisms of discrimination against marginalized groups.

The idea that health-equity research is a mechanism to develop and implement interventions to attain health equity generated strong feelings of inadequacy. This led to discussions about measurement and methodology. For example, some were uncomfortable with the idea of measuring racism and discrimination but were more comfortable with the quantitative idea of measuring representation, although they acknowledged that lack of representation may be driven by racism and discrimination.

Some participants faced major barriers and frustrations and noted specific and critical needs and lack of support for investigators in this area, from idea to funding to project completion and publication. Concerns included the structural biases that decrease the perceived value of this work; the inherent career costs relative to other pathways in academic medicine; the important but under-recognized value of advocacy in general; and advocacy’s lack of a place in standard notions of scholarship, career advancement, and promotion.

Participants discussed the costs for faculty members from marginalized groups, including undue expectations to support institutional DEIA efforts, which further exacerbates the burden of research in this difficult area. One of the participants noted the gender representation of the focus groups and observed: **“It’s actually a national issue when we look at how these priorities and health-equity research or social-justice work is generally led by, most often, women and not men. So that’s another area that we need to advocate for, because when you look at who are the directors of [research] centers is generally men…”**

Despite the stated barriers, all participants felt that this area needs to be addressed and that doing so would benefit patients and the institutions that serve them.

Participants identified the need for intentional and positive ongoing engagement with marginalized communities and community partners.

Participants also recognized the need for a strong community of investigators who are committed to this topic; will incorporate health equity into their current research program; will engage experts in the field; and will collaborate with, mentor, and support those who are seeking to pursue this work.

Overall, the participants expressed interest and commitment to this area of research and desire for further engagement and support.

## DISCUSSION

### Increasing diversity in Vermont and in the healthcare workforce

Vermont’s population exhibits increases in racial and other elements of diversity. In highly populated areas, such as Chittenden county there has been an increase, in numbers and proportions, of Hispanic, Black/African, Aboriginal, Native Hawaiian/islander, and multiracial peoples (https://datausa.io/profile/geo/chittenden-county-vt#demographics) (data and analyses available on request).

### Health-care workforce diversity data from Vermont

We have observed similar trends in diversity within the healthcare workforce through an analysis of census data of Vermont providers. Only since 2014 has there been data allowing estimation of the diversity of the state’s healthcare workforce (Courtesy Jessica Moore; data and analyses available on request). However, the relevant retention rate is unmeasured and thus not known^12^.

Growing a diverse academic workforce will need enhanced mechanisms of recruitment and retention. An important retention factor is success in scholarly activities. The success rate of federal funding by investigators in marginalized groups is less than that in white-identified groups^23^. While this is likely related to structural racism in academic medicine, one clearly defined factor is that faculty of color seek to do research in areas relevant to health equity and care of marginalized populations. This is less-well funded than other areas^23^ and may contribute to the overall decreased success^24^.Therefore, an increase in federal sources of support for such research could increase the diversity of academic faculty, provided it does not solely incentivize majority investigators to seek additional funding for their ongoing research programs. To diversify, academic institutions in states such as Vermont must provide financial support for this work in addition to building other resources to further support, embed, and center diversity and benefit faculty of color and from marginalized groups. Even incrementally enhanced support may yield significant productivity, portfolio, and engaged participants.

### Redefinition and revitalization

Much existing research focuses on health disparities driven by rurality^25^ and reduced access to care, patient behavior, or race as denoting genetic predisposition, with limited assessment of social determinants and societal mechanisms of marginalization. Few Vermont investigators focus on the intricate role potentially played by systemic factors in health inequity. However, Vermont’s academic medical center is affiliated with a larger academic community with researchers focused on issues such as environmental and criminal justice and food systems, which may be incorporated into the examination of Health Equity.

## RECOMMENDATIONS

### Further work is needed to

1. Identify and implement appropriate guidelines, “roadmaps” and strategies to improve health equity research
2. Refocus and revitalize existing infrastructure to build robust, integrated capacity in health equity research
3. Apply the highest standards of cultural and linguistic guidelines to the existing IRB structure.
4. Building on work already initiated through collaborations between the UVM Office of Diversity and the UVM Medical Center, create additional mechanisms to connect the Health Equity research community, including information clearinghouses, websites and opportunities for round table discussions.
5. Increase direct support for Health Equity research through internal funding, assignment of administrative (e.g., pre and post grant award) resources, and building dedicated infrastructure with specific funding allocated to support Health Equity research initiatives.
6. Align institutional funding and financial priorities with Health Equity

## CONCLUSION

A move towards health equity would enable Vermont health educators and providers to facilitate conditions wherein equity permeates every process and workflow, including biomedical research. While there are no simple one-size-fits-all solutions, there are specific and measurable goals that Vermont scholars, educators and practitioners can take to move towards health equity. Considering these issues for all Vermonters has never been more important.

## Data Availability

All data produced in the present work are contained in the manuscript

## ACKNOWLEDGMENTS

The authors would like to acknowledge that the University of Vermont *sits within a place of gathering and exchange, shaped by water and stewarded by ongoing generations of Indigenous peoples, in particular the Western Abenaki*.

We also acknowledge support from the Executive Leadership in Academic Medicine (ELAM) program (to EAB), the LCOM Equity & Inclusion Excellence Certificate Cohort program (to GB), and to the University of Vermont Office of the Vice President for Research, Departments of Obstetrics, Gynecology and Reproductive Sciences and Pediatrics and Vermont Leadership Education in Neurodevelopmental Disabilities Program.

Transcription Services were provided by Maria Clara Avila, RN, MS.

